# Cost-effectiveness of paediatric influenza vaccination in the Netherlands

**DOI:** 10.1101/2020.03.02.20029124

**Authors:** Pieter T. de Boer, Franklin C.K. Dolk, Lisa Nagy, Jan C. Wilschut, Richard Pitman, Maarten J. Postma

## Abstract

**Background:** This study evaluates the cost-effectiveness of extending the Dutch influenza vaccination programme for elderly and clinical risk groups to include paediatric influenza vaccination, taking indirect protection into account.

**Methods:** An age-structured dynamic transmission model was used that was calibrated to influenza-associated GP visits over four seasons (2010/11 to 2013/14). The clinical and economic impact of different paediatric vaccination strategies were compared over 20 years, varying the targeted age range, the vaccine type for children and the vaccine type for elderly and clinical risk groups. Outcome measures include averted symptomatic infections and deaths, societal costs and quality-adjusted life years (QALYs), incremental cost-effectiveness ratios, and net health benefits (NHBs), using a willingness-to-pay threshold of €20,000 per QALY gained.

**Results:** At an assumed coverage of 50%, adding vaccination of 2- to 17-year-olds with quadrivalent-live-attenuated influenza vaccine (Q-LAIV) to the current influenza vaccination programme was estimated to avert on average 406,270 symptomatic cases and 83 deaths per season compared to vaccination of elderly and risk groups with trivalent inactivated vaccine (TIV), and was cost-saving (cumulative 20-year savings of 36,396 QALYs and €1,680 million; NHB: 120,411 QALYs). This strategy dominated paediatric vaccination strategies targeting 2- to 6-year-olds or 2- to 12-year-olds, or paediatric vaccination strategies with TIV. The highest NHB was obtained when 2- to 17-year-olds were vaccinated with Q-LAIV and existing target groups switched from TIV to quadrivalent inactivated vaccine (NHB: 132,907 QALYs).

**Conclusion:** Modelling indicates that paediatric influenza vaccination reduces the disease burden of influenza substantially and is cost-saving.

## 1. Introduction

Seasonal influenza epidemics are responsible for a significant health and economic burden.^1,2^ Although the most severe outcomes occur among elderly and persons with chronic diseases, increasing evidence shows that the burden of influenza among children is also relevant.^3^ Young children are frequently hospitalized, require an outpatient visit, or stay at home from school, causing work loss among caregivers.^4,5^ Furthermore, children are thought to play a key role in the transmission of influenza because they remain infectious for a longer period and have more close contacts than adults.^6^ Ecological studies as well as mathematical modelling studies suggest that paediatric influenza vaccination would provide not only direct protection but also indirect protection to susceptible contacts due to herd immunity.^7-12^

Anticipating these direct and indirect benefits, several countries have issued positive recommendations for vaccination of children against influenza.^13^ For instance, the United Kingdom is currently rolling out a publicly funded influenza vaccination programme for children aged 2-16 years using the intranasally administered quadrivalent live-attenuated influenza vaccine (Q-LAIV).^14^ In the Netherlands, influenza vaccination is offered free of charge to all individuals aged ≥60 years and individuals aged <60 years with certain chronic diseases.^15^ In 2007, vaccination of healthy children against influenza was not recommended by the Health Council of the Netherlands, as the risk of severe complications and mortality among children was not considered high enough.^16^ However, the discussion went ongoing and the decision on paediatric influenza vaccination will be reassessed by the Health Council of the Netherlands in 2020.^17^

Cost-effectiveness is an important aspect in the decision framework for the implementation of vaccination programmes in most countries including the Netherlands.^18^ Against this perspective, we conducted a cost-effectiveness analysis of inclusion of paediatric influenza vaccination in the current vaccination programme for elderly and clinical risk groups in the Netherlands. As paediatric influenza vaccination is expected to confer indirect effects upon the wider community, a dynamic transmission model was used that accounts for herd immunity.

## 2. Methods

### 2.1. Overview

The analysis uses a probabilistic sensitivity analysis (PSA) approach, taking into account the uncertainty in the transmission, clinical, and economic parameters. A deterministic transmission model was used to simulate the population-level dynamics of influenza infection (Section 2.2). To incorporate uncertainty in the transmission parameters, a set of key transmission parameters was repeatedly sampled from input distributions. Those sets that fitted the observed information from the Netherlands were retained and are collectively referred to as the ‘calibrated model’ (section 2.3). The updated sets of parameter distributions were then integrated with the transmission model to produce a PSA of the transmission parameter inputs. The results of the transmission model served as an input for the economic PSA in which the clinical and economic parameters were sampled and outcomes were compared for a range of vaccination policies (section 2.4)

### 2.2. Dynamic transmission model

The dynamic transmission model is a compartmental model, stratified by age in months. The model uses a SEIR_F_R_L_S(V) structure (susceptible, exposed, infectious, recovered with short-term immunity, recovered with long-term immunity, vaccinated). Ageing was simulated on a monthly basis, informed by Dutch data on age structure, birth rates, and mortality rates.^19-21^ To emulate the observed influenza dynamics, the model population was seeded annually with new infectious influenza cases. Contact rates between age groups were obtained from an age-stratified mixing matrix from the section of the POLYMOD study specific for the Netherlands.^22^ The magnitude of these age-specific contact rates and the probability of transmission per contact yielded an age-stratified matrix of transmission coefficients. As the incidence of influenza follows a marked seasonal pattern, the magnitude of these transmission coefficients was assumed to vary sinusoidally over time, peaking near the end of the calendar year.

Influenza A and influenza B were simulated independently. Within an influenza type, two strains were assumed to be modelled: a strain from each of the H1N1 and H3N2 subtypes for influenza A, and a strain from each of the Victoria and Yamagata lineages for influenza B. Both strains were modelled simultaneously, so the model compartmental structure combined the SEIR_F_R_L_S(V) structure of each. No cross-protective immunity between strains was assumed. The proportion of the population immune, by age and virus, at the model start was estimated by running the model forward, then obtaining the compartmental populations from a year in which the model incidence approximated the observed incidence of the first season of the calibration period.^23^

The vaccination campaign was assumed to start in mid-October. During the calibration period, the model simulated vaccination using trivalent inactivated vaccine (TIV) with the vaccine composition as recommended by the WHO.^24^ Influenza vaccine uptake for 2010/2011 to 2013/14 was obtained from the literature.^25^ Efficacy against laboratory-confirmed influenza was applied by age.^26,27^ No cross-protection of TIV against the non-included influenza B lineage was assumed. The duration of vaccine-induced protection was assumed to be much shorter, on average, than that of naturally acquired immunity.

More details are provided in the Supplementary Methods.

### 2.3. Calibration

In the calibration stage, the transmission model was run for the 2010/11 through 2013/14 seasons for each set of input parameter samples. The main data source for the model calibration was a set of Netherlands-specific general practitioner (GP) consultation rates obtained from a regression of influenza-like-illness (ILI) consultation data against laboratory-confirmed influenza reports.^23^ Influenza-associated GP consultation rates were stratified by influenza strain, age group (0-4, 5-19, 20-59, and ≥60 years), and season (2010/11-2013/14). Prior to each simulation in the model calibration stage, the transmission parameters were sampled. We used subtype/lineage-specific consultation rates from the regression analysis in which the unspecified influenza-positive samples were not redistributed to influenza subtype/lineage. The resulting simulated influenza GP consultation of each simulation was compared with observed data from the same period using the Poisson deviance and a set of other fit criteria (Supplementary File 1). Sets of parameter samples that met the criteria were retained for the calibrated model (Table 1 and Supplementary File 1).

**Table 1:** Key transmission model inputs

| Input | Stratified by | Distribution | Min | Max |
| --- | --- | --- | --- | --- |
| **R0 parameters** |  |  |  |  |
| Transmission coefficient | Virus | Uniform | 2.76E-08 | 8.29E-08 |
| Latent period (days) | Virus | Uniform | 0.01 | 3 |
| Infectious period (days) | Virus | Uniform | 0.5 | 5 |
| **Immunity parameters** |  |  |  |  |
| Duration of initial naturally acquired immunity (years) | Virus | Uniform | 0.5 | Influenza A: 10<br>Influenza B: 30 |
| Duration of long-term naturally acquired immunity (years) | Virus | Uniform | 10 | 70 |
| Probability of acquiring long-term immunity | Virus | Uniform | 0 | 1 |
| Duration of vaccine-induced immunity (years) | Virus | Uniform | 0.5 | 3 |
| **Vaccination parameters** |  |  |  |  |
| Vaccine efficacy |  |  |  |  |
| 0-17y |  | Lognormal: 48<br>(95%CI: 31-61) |  |  |
| 18-64y |  | Lognormal: 59<br>(95%CI: 50-66) |  |  |
| ≥65y |  | Lognormal: 50<br>(95%CI: 39-59) |  |  |
| Vaccination campaign duration |  | Uniform (integer) | 30 | 40 |
| **Probabilities** |  |  |  |  |
| Probability of symptoms given infection |  | Beta: Mean = 0.669,<br>SE = 0.0413 | 0 | 1 |

R0: Basic reproduction number, SE: Standard error

### 2.4. Expected net benefit analysis

The calibrated model was used to compare the clinical and economic impact for a range of vaccination strategies. For each set of parameter samples of the calibration, the model was run forward from 2010/11 to 2034/35. Explored vaccination strategies diverged from the 2015/16 season, and results from the period 2015/16 to 2034/35 were used for the analysis (time-horizon of 20 years). The initial output of the model integration concerned incidence of infection. Risk functions of clinical outcomes were applied to the outcomes of the transmission model in order to estimate the number of symptomatic cases, GP visits, hospitalizations, and deaths. Estimates of costs and quality-adjusted life years (QALYs) lost were then applied to the clinical outcomes. Costs were discounted at 4% per year and QALYs at 1.5% per year from the start of the 2015/16 season.^28^

#### 2.4.1. Vaccination strategies

Explored vaccination strategies are listed in Table 2. For the existing vaccination programme for elderly and risk groups, annual vaccination with TIV or quadrivalent inactivated vaccine (QIV) were considered. QIV includes an additional influenza B virus lineage compared with TIV. A strategy of no influenza vaccination at all was also added.

**Table 2:** Explored vaccination strategies.

| Scenario name | Description |
| --- | --- |
| No vaccination | No influenza vaccination in any age |
| TIV | TIV in individuals 6 months of age and older |
| QIV | QIV in individuals 6 months of age and older |
| TIV/TIV | Paediatric programme<sup>a</sup> with TIV; TIV in other age groups. |
| TIV/Q-LAIV | Paediatric programme<sup>a</sup> with Q-LAIV; TIV in other age groups. |
| QIV/Q-LAIV | Paediatric programme<sup>a</sup> with Q-LAIV; QIV in other age groups. |

QIV: Quadrivalent inactivated influenza vaccine, Q-LAIV: Quadrivalent live-attenuated influenza vaccine, TIV: Trivalent inactivated influenza vaccine.
<sup>a</sup> Paediatric programmes were explored for the age groups 2–6, 2–12 or 2–17 years. The main analysis assumes a vaccination coverage in healthy children of 50%.

For the additional paediatric vaccination strategies, annual vaccination with TIV and Q-LAIV were considered. As post-licensure studies comparing the effectiveness of LAIV with inactivated vaccine (IV) found equivocal results,^29-31^ the efficacy of LAIV was assumed to be equal to that of IV. Hence, clinical outcomes for Q-LAIV also represent those of QIV. Vaccination was considered for the age groups 2-6 years, 2-12 years, and 2-17 years. The paediatric vaccination programme was assumed to be introduced for the whole age range at the same time and children received only one dose irrespective of whether influenza vaccine had been received before.

The vaccination coverage of the paediatric programme was assumed at 50%, in accordance with emerging UK data on uptake during a paediatric vaccination programme.^32^ In age groups outside the paediatric vaccination programme, the vaccine uptake was unchanged and the latest data (2013/14 season) were carried forward.

#### 2.4.2. Clinical outcome risk functions

The probability of symptoms, given influenza infection, was obtained from the literature.^33^ Age-specific probabilities of a GP consultation, given infection, was calculated as part of the calibration using the GP consultation rates and the modelled incidence of infection, in order to calculate the deviance of the run compared with the GP rate data.^23^ Age-specific probabilities of hospitalization were based on Dutch estimates of the relationship between respiratory-associated hospitalization and ILI incidence at the GP.^34^ Age-specific probabilities of death were based on Dutch incidence rates of respiratory-associated influenza death.^35^ More details are provided in Supplementary File 1.

#### 2.4.3. Economic outcomes

The economic analysis adopted a societal perspective. All costs were converted to 2017 euros using the Dutch consumer price index.^36^ We distinguished healthcare costs (vaccine, administration, GP visits including prescribed medication and specialist visits, hospitalizations, and healthcare costs unrelated to influenza in gained life years [indirect healthcare costs]), patient costs (over-the-counter medication and travel costs), and productivity losses (from patients or caregivers of sick children). The tendered vaccine price of TIV in the Netherlands was €3.59 in the 2017/18 season (latest available at time of calculations).^37^ The vaccine price of QIV was assumed to be 50% higher than that of TIV and the vaccine price of Q-LAIV was assumed to be equal to that of QIV. Since the vaccine efficacy of Q-LAIV and QIV were also assumed to be equal, economic results of Q-LAIV represent also those of QIV. Costs of vaccine administration and influenza disease were obtained from published sources or national datasets.

QALYs lost due to influenza illness was based on published studies that used the validated EuroQol-5 Dimensions (EQ-5D) instrument.^38^ QALYs lost due to premature death were calculated using age-specific life expectancies at age of death and Dutch population norms for quality of life.^39^ To account for the increasing life expectancy over time, life expectancy predictions from Statistics Netherlands of the year 2024 (halfway through the analysed time horizon) were used.^40^

More details are presented in Supplementary File 1.

#### 2.4.4. Cost-effectiveness

The base-case estimate per vaccination scenario was obtained by averaging the clinical and economic results across simulations. The incremental cost-effectiveness ratio (ICER) was calculated by dividing the difference in costs by the difference in QALYs. Results are also presented as net health benefits (NHB), converting monetary outcomes into QALYs using a willingness-to-pay threshold λ (in €per QALY). The NHB is calculated as ΔQALY – (ΔCosts/λ), and a positive NHB indicates that the intervention is cost-effective to λ. We used a λ of €20,000 per QALY gained, which is the lowest Dutch threshold published, and is often applied for prevention programmes.^41^ Cost-effectiveness acceptability curves (CEACs) are drawn to present the probability of having the highest NMB over a range of cost-effectiveness thresholds. As the policy with the highest NHB may not always be the optimal decision (for instance, a policy with the highest NHB could be subject to extended dominance^42^), the probability of optimum policy was shown in a cost-effectiveness acceptability frontier (CEAF).

#### 2.4.5. Univariate sensitivity analysis

Univariate sensitivity analyses were performed to test for structural uncertainty, including variation of the vaccination coverage, a higher efficacy of LAIV based on clinical trial data,^43^ and the vaccine price of Q-LAIV.

## 3. Results

### 3.1. Calibration

During the calibration stage, 7,198 simulations were selected as a close enough fit to the Dutch data. The resulting updated distributions are plotted in the Supplementary Results (Supplementary File 2: Figure S2.1-Figure S2.7 and Table S2.1-Table S2.3), as are visual comparisons of the model incidence to the GP regression data (Supplementary File 2: Figure S2.8-Figure S2.11). Sampling from uniform input distributions, and keeping the samples that met the calibration criteria, produced clearly defined unimodal distributions for the basic reproduction number; these distributions were clearly updated from their initial inputs by the acceptance-rejection sampling according to the calibration heuristic (Supplementary File 2: Figure S2.2).

### 3.2. Clinical impact

Table 3 shows the expected 20-year average seasonal number of clinical events in the Netherlands for various vaccination scenarios (outcomes of other strategies are available in Supplementary File 2: Table S2.4). Across 7,198 simulations are presented, vaccination of elderly and risk groups with TIV was estimated to prevent on average 202,931 (95% range: 69,058-522,523) symptomatic cases, 51,698 (19,725-112,215) GP visits, 1,134 (467-2,331) hospitalizations and 249 (101-582) deaths per season compared with no influenza vaccination.

**Table 3:** 20-year average seasonal number of clinical outcomes in the Netherlands. For paediatric vaccination strategies, vaccination coverage in children was assumed at 50%.

| Scenario | Symptomatic infections |  |  | GP visits |  |  | Hospitalizations |  |  | Deaths |  |  |
| --- | --- | --- | --- | --- | --- | --- | --- | --- | --- | --- | --- | --- |
|  | Exp. | (95% range)<sup>a</sup> | Rate<sup>b</sup> | Exp. | (95% range)<sup>a</sup> | Rate<sup>b</sup> | Exp. | (95% range)<sup>a</sup> | Rate<sup>b</sup> | Exp. | (95% range)<sup>a</sup> | Rate<sup>b</sup> |
| No vaccination | 1,157,285 | (510,878–4,186,700) | 6,755 | 172,603 | (85,867–425,223) | 1,007 | 2,560 | (1,073–7,295) | 14.9 | 523 | (153–2,377) | 3.05 |
| TIV | 954,353 | (462,235–2,960,578) | 5,570 | 120,906 | (71,688–212,412) | 706 | 1,426 | (700–2,961) | 8.32 | 274 | (76–1,152) | 1.60 |
| QIV | 898,133 | (445,978–2,848,591) | 5,242 | 114,729 | (68,743–201,573) | 670 | 1,292 | (638–2,451) | 7.54 | 213 | (57–941) | 1.24 |
| TIV/TIV in 2–6y | 853,115 | (362,641–2,706,864) | 4,979 | 106,125 | (46,046–192,326) | 619 | 1,274 | (582–2,730) | 7.44 | 264 | (74–1,129) | 1.54 |
| TIV/TIV in 2–12y | 734,350 | (225,422–2,505,900) | 4,286 | 86,626 | (23,822–187,957) | 506 | 1,086 | (302–2,767) | 6.34 | 250 | (60–1,091) | 1.46 |
| TIV/TIV in 2–17y | 691,051 | (203,290–2,351,643) | 4,033 | 79,320 | (23,797–183,698) | 463 | 1,006 | (272–2,721) | 5.87 | 240 | (56–1,070) | 1.40 |
| TIV/Q-LAIV in 2–17y | 548,084 | (74,821–2,200,169) | 3,199 | 67,308 | (7,856–134,861) | 393 | 843 | (108–1,979) | 4.92 | 191 | (19–878) | 1.12 |
| QIV/Q-LAIV in 2–17y | 496,313 | (26,593–2,075,552) | 2,897 | 61,972 | (3,188–126,244) | 362 | 733 | (35–1,695) | 4.28 | 141 | (5–814) | 0.825 |

GP: General practitioner, QIV: Quadrivalent inactivated vaccine, Q-LAIV: Quadrivalent live-attenuated influenza vaccine, TIV: Trivalent inactivated vaccine, y: years of age.
<sup>a</sup>:Exp: Expectation (average over 7,198 simulations) with range in which 95% of simulations fall. <sup>b</sup>: Rate per 100,000 person years.

Introducing paediatric vaccination at an assumed coverage of 50% was estimated to prevent a substantial additional number of clinical events, and its impact increases by targeting a broader age group or by using Q-LAIV instead of TIV or both. Adding TIV for 2- to 17-year-olds was estimated to prevent on average 263,302 (95% range: 108,775-598,074) symptomatic cases, 41,585 (15,539-84,524) GP visits, 420 (118-937) hospitalizations, and 34 (-17-116) deaths per season compared with TIV for elderly and risk groups. Adding Q-LAIV for 2- to 17-year-olds was estimated to prevent on average 406,270 (95% range: 225,122-775,282) symptomatic cases, 53,597 (24,625-100,130) GP visits, 583 (252-1,156) hospitalizations, and 83 (30-180) deaths per season compared with TIV for elderly and risk groups. The lowest number of clinical events was estimated for the use of QIV for elderly and risk groups and Q-LAIV for 2- to 17-year-olds.

Herd immunity amongst other age groups contributed substantially to this reduction. For adding Q-LAIV for 2- to 17-year-olds, 50% of all prevented symptomatic cases and 99% of all prevented deaths were in other age-groups than 2- to 17-year-olds (see Supplementary File 2: Figure S2.12).

Paediatric vaccination induced an age-shift of influenza cases to older age groups. For instance, introducing vaccination for 2- to 6-year-olds increased the number of symptomatic cases among 10- to 17-years-olds (Supplementary File 2: Figure S2.12).

### 3.3. Cost-effectiveness

Table 4 summarizes the expected 20-year cumulative total costs and QALY loss for various vaccination scenarios (outcomes of other strategies are available in Supplementary File 2: Table S2.5). Compared with no vaccination, TIV for elderly and risk groups was estimated to save 35,627 QALYs and to cost €63 million, resulting in an ICER of €1,776/QALY gained. A switch from TIV to QIV for the elderly and risk groups gave an estimated further gain of QALYs, while saving costs—i.e., QIV dominated TIV.

**Table 4:** 20-year cumulative economic outcomes. Vaccination coverage in children in paediatric vaccination strategies was assumed at 50%. Results include an annual discount rate of 4% for costs and 1.5% for QALYs.

| Policy | Total QALYs<br>(thousands) | Total costs<br>(€, millions) | QALY gain<br>(thousands)<br><sup>a</sup> | Costs<br>(millions)<sup>a</sup> | ICER (€/QALY<br>gained)<sup>b</sup> | NHB<br>(thousands)<sup>a,c</sup> |
| --- | --- | --- | --- | --- | --- | --- |
| No vaccination | 125.41 | 6,687 |  |  |  | (32.5) |
| TIV | 89.78 | 6,750 | 35.63 | 63 | 1,776 | - |
| TIV/TIV in 2–6y | 81.45 | 6,293 |  |  | Dominated | 31.2 |
| QIV | 80.87 | 6,659 |  |  | Dominated | 13.5 |
| TIV/Q-LAIV in 2–6y | 77.78 | 6,142 |  |  | Dominated | 42.4 |
| TIV/TIV in 2–12y | 71.58 | 5,788 |  |  | Dominated | 66.3 |
| QIV/ Q-LAIV in 2–6y | 69.08 | 6,041 |  |  | Dominated | 56.2 |
| TIV/TIV in 2–17y | 67.73 | 5,658 |  |  | Dominated | 76.7 |
| TIV/ Q-LAIV in 2–12y | 59.93 | 5,298 |  |  | Dominated | 102.5 |
| TIV/ Q-LAIV in 2–17y | 53.38 | 5,070 |  |  | Dominated | 120.4 |
| QIV/ Q-LAIV in 2–12y | 51.90 | 5,195 |  |  | Dominated | 115.7 |
| QIV/ Q-LAIV in 2–17y | 45.80 | 4,972 | 43.98 | -1,779 | Cost-saving | 132.9 |

ICER: Incremental cost-effectiveness ratio, NHB: Net health benefit, QALY: Quality-adjusted life year, QIV: Quadrivalent inactivated vaccine, Q-LAIV: Quadrivalent live-attenuated influenza vaccine, Trivalent inactivated vaccine, y: years of age. <sup>a</sup>: Compared with vaccination with TIV at current uptake rates. <sup>b</sup>: Vaccination policies were listed as dominated when there was another policy with a QALY gain against lower costs (strict dominance) or a QALY gain against a lower ICER (extended dominance). <sup>c</sup>: Calculated as: $QALY - (Cost / \lambda)$ , with $\lambda = €20,000/QALY$ .

All paediatric vaccination scenarios were expected to dominate the current strategy of vaccination of elderly and risk groups with TIV, and each extension of the targeted paediatric age group and/or a switch from TIV to Q-LAIV dominated the preceding scenario. For instance, adding TIV for 2- to 17-year-olds resulted in 20-year cumulative savings of 22,050 QALYs and €1,092 million (NHB: 76,665 QALYs), while adding Q-LAIV for 2- to 17-year-olds would save 36,396 QALYs and €1,680 million (NHB: 120,411 QALYs). Most paediatric vaccination strategies dominated a switch from TIV to QIV for elderly and risk groups. Considering all strategies, vaccination of elderly and risk groups with QIV and 2- to 17-year-olds with Q-LAIV dominated all other scenarios. The 20-year cumulative savings of this strategy were estimated at 43,977 QALYs and €1,779 million (NHB: 132,907 QALYs) compared with TIV for elderly and risk groups.

The uncertainty around the economic impact of paediatric vaccination was considerable (Figure 1A). The 95% ranges of adding Q-LAIV for 2- to 17-year-olds were 19,383-69,473 QALYs and €693 million-€3,736 million compared with TIV for elderly and risk groups, resulting in a 95% range of the NHB of 54,032-256,268. All simulations for this strategy resulted in a gain of QALY and lower costs. For adding TIV for 2- to 6-year-olds, however, 2.3% of the simulations resulted in an overall QALY loss (Supplementary File 2: Table S2.7).

**Figure 1:**
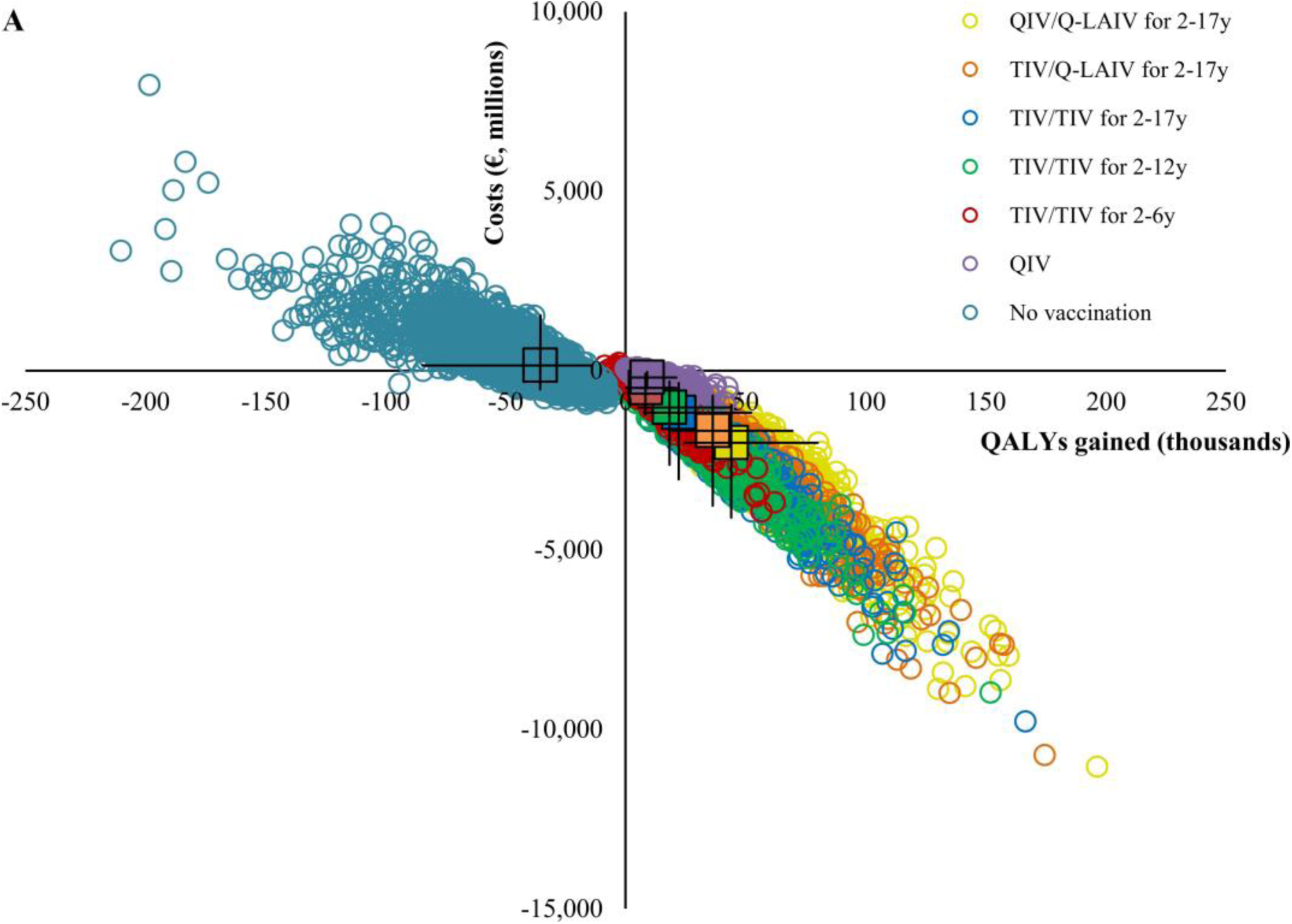
Results of a probabilistic sensitivity analysis (7,198 simulations) of a selection of influenza vaccination strategies in the Netherlands over 20 seasons. Vaccination coverage in children was assumed at 50%. A) The cost-effectiveness plane shows the incremental costs and incremental QALYs of different vaccination strategies compared with TIV for risk groups. A square represents the average across simulations and bars represent the range in which 95% of the simulations fell. Only a selection of scenarios were presented to enhance the visibility of the figure.

**Figure 1:**
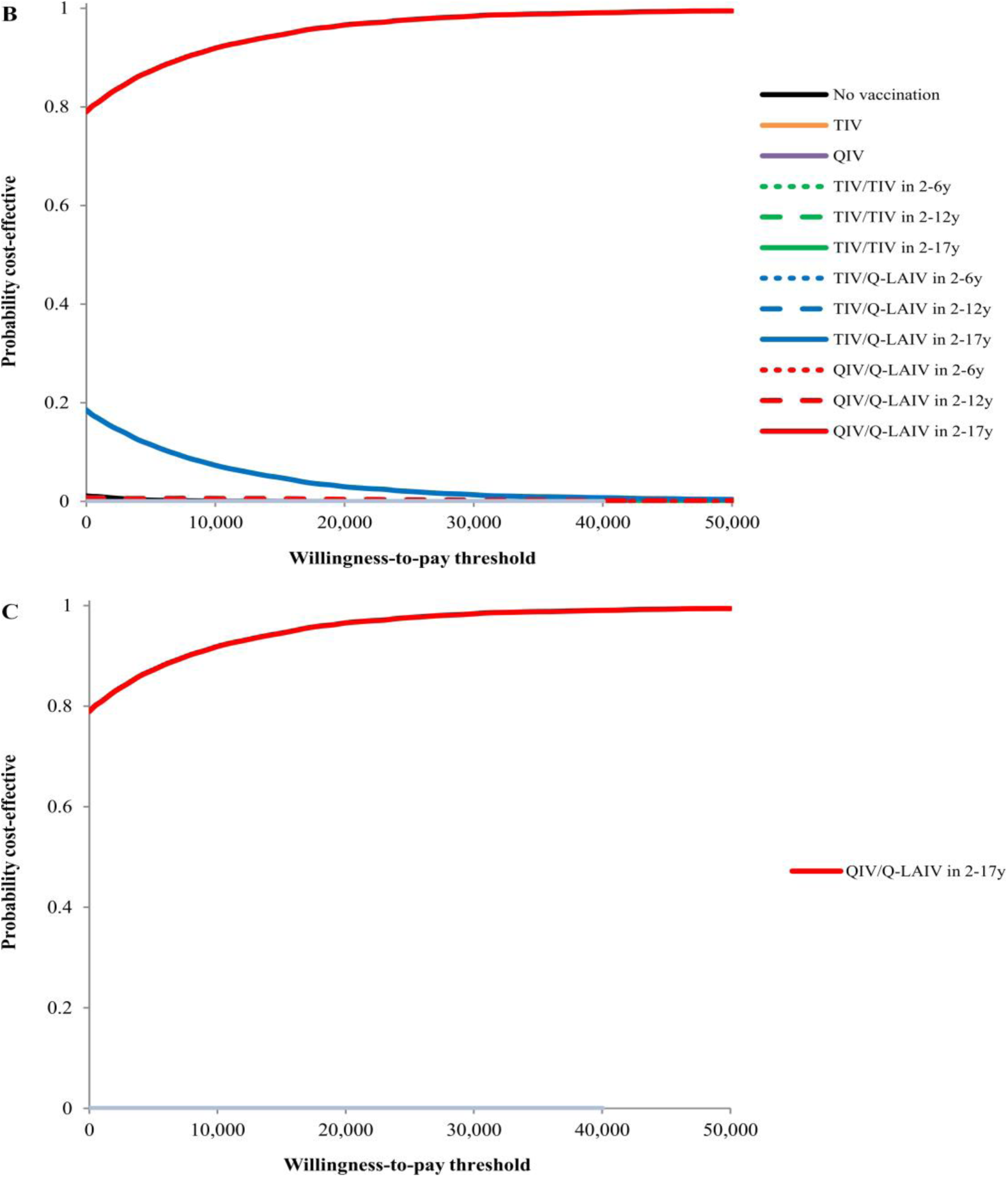
Results of a probabilistic sensitivity analysis (7,198 simulations) of a selection of influenza vaccination strategies in the Netherlands over 20 seasons. Vaccination coverage in children was assumed at 50%. B) The cost-effectiveness acceptability curve shows the probability of having the highest NHB for a selection of vaccination strategies. C) The cost-effectiveness acceptability frontier shows the probability of being the most cost-effective alternative for a selection of vaccination strategies. QALY: Quality-adjusted life year, QIV: Quadrivalent inactivated vaccine, Q-LAIV: Quadrivalent live-attenuated influenza vaccine, TIV: Trivalent inactivated vaccine, y: years of age.

The CEAF indicates that QIV for elderly and risk groups and Q-LAIV for 2- to 17-year-olds also had the highest probability of being the optimum policy at any willingness-to-pay threshold considered (Figure 1B and C).

### 3.4. Univariate sensitivity analysis

Figure 2 shows the univariate sensitivity analysis of extending TIV for elderly and risk groups with Q-LAIV for 2- to 17-year-olds. Varying the vaccination coverage in children between 20% and 80% indicated a non-linear relationship between the coverage and the NHB, with increasing coverage resulting in relatively lower returns. Nevertheless, paediatric vaccination strategies at 80% coverage dominated strategies at 20% or 50% coverage (Supplementary File 2: Table S2.5-Table S2.6). Using a vaccine efficacy of Q-LAIV based on trial data increased the average prevented number of symptomatic infections from 406,270 to 566,465 (Supplementary File 2: Table S2.4) and the NHB from 120,411 to 175,263 QALYs (Supplementary File 2: Table S2.8). Applying a higher QALY loss for influenza illness also had a high impact on the NHB, as the majority of QALYs gained were due to prevention of influenza illness rather than prevention of influenza deaths (Supplementary File 2: Figure S2.13A), The cost-effectiveness of the paediatric vaccination was not sensitive to the vaccine price. The productivity losses of influenza represented the majority of the cost burden of influenza (Supplementary File 2: Figure S2.13B) and adopting a healthcare payer’s perspective resulted in an ICER of €13,132/QALY gained (Supplementary File 2: Table S2.8) and an NHB of 13,491 QALYs.

**Figure 2:**
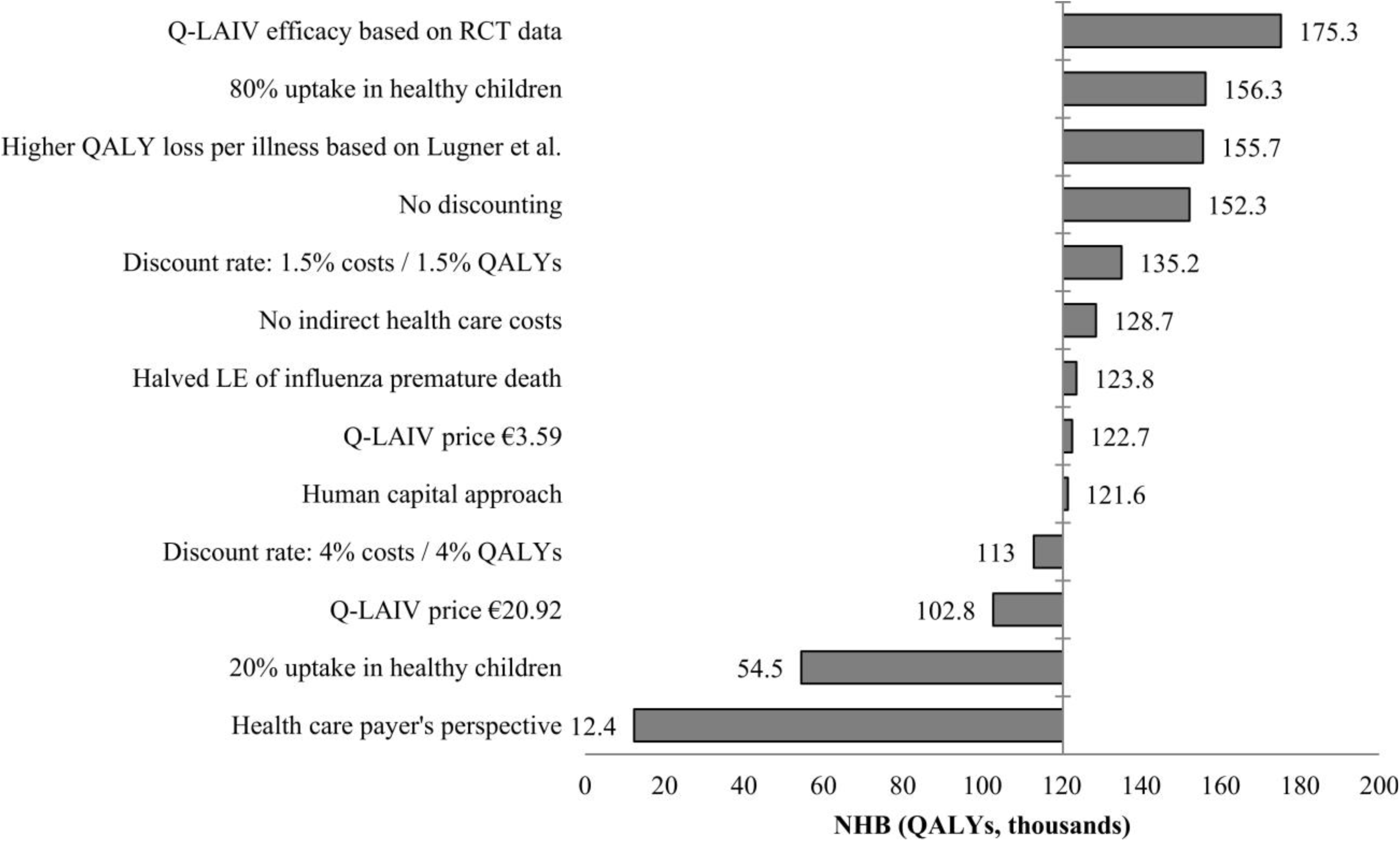
Scenario analysis of paediatric influenza vaccination of 2- to 17-year-olds with quadrivalent live-attenuated influenza vaccine, while other target groups were vaccinated with trivalent inactivated vaccine. The base case analysis assumes a vaccination coverage in children of 50%. Cost-effectiveness results are summed up over a time horizon of 20 years. RCT: Randomized clinical trial, QALY: Quality-adjusted life year, Q-LAIV: Quadrivalent live-attenuated vaccine.

### 3.5. Direct impact

Supplementary file 2: Table S2.11 shows the economic results when only costs and effects in 2- to 17-year-olds themselves are considered. Adding Q-LAIV for 2- to 12-year-olds at 50% coverage was found to dominate the current strategy of TIV for risk groups (20-year cumulative savings of €20 million and 12,138 QALYs; NHB 13,137 QALYs), and dominated also other paediatric strategies considered. Extending vaccination with Q-LAIV from 2- to 12-year-olds to 2- to 17-year-olds would cost €30,470 per QALY gained.

## 4. Discussion

Introduction of childhood influenza vaccination was estimated to prevent substantial morbidity and mortality on a population-level, and to be cost-saving from a societal perspective. The NHB increased when a broader paediatric age range was targeted or when Q-LAIV was used instead of TIV. The highest NHB obtained with the vaccination of 2- to 17-year-olds with Q-LAIV and elderly and risk groups with QIV.

Indirect protection made a pronounced contribution to the cost-effectiveness of paediatric vaccination, given that half of the prevented symptomatic cases and nearly all prevented deaths were among adults. This reduction in mortality occurred despite the already high vaccination coverages among elderly. Adding paediatric vaccination was also estimated to be cost-saving when elderly and risk groups would be vaccinated with QIV (TIV is replaced by QIV for the 2019-20 season in the Netherlands).^44^ Furthermore, adding paediatric vaccination was also estimated to be cost-saving when only costs and effects among children themselves are considered.

There was a non-linear relationship between coverage and effects of vaccination. This is explained by the concept that once a critical uptake rate has been achieved, further increase yields diminishing returns.^45^ Nonetheless, the economic returns of paediatric vaccination were estimated to be such that the NHB kept increasing with increasing coverage. Paediatric vaccination is also likely to induce an age-shift of influenza infections to older age groups. This age-shift occurs because the probability of becoming infected is lower; hence, infection occurs at an, on average, older age. However, this age-shift did not outweigh the benefits of paediatric vaccination as a whole, and could also be considered as a good thing, as there is a lowering of the likelihood of infection in the very young.

As recommended by international guidelines,^46,47^ a dynamic transmission model was used that accounts for indirect effects of vaccination and that captures the gains and losses of immunity over time. The transmission model was calibrated using Dutch influenza epidemiology data for four seasons in order to obtain realistic parameter sets, and we used Dutch data on contact patterns, outcome probabilities, and economic parameters.

The waning rate of immunity and vaccine efficacy was assumed to remain constant over time; however, these parameters may vary between seasons due to irregular antigenic drift and vaccine match. A recent modelling study that accounted for seasonal variation estimated that paediatric vaccination may increase the variability in the scale of the epidemic; that is, seasons with small epidemics are occasionally alternated with seasons with very large epidemics due to a build-up of susceptibles.^48,49^ Increased variability in epidemic size could potentially lead to less impact of the paediatric vaccination programme or a small risk of an overall QALY loss.^50^

There is ongoing debate about the vaccine effectiveness of LAIV. Clinical trials found that the efficacy of LAIV was superior to that of IVs,^26^ whereas effectiveness studies found the effectiveness of LAIV to be superior, similar, or inferior to IV.^29-31^ Use of vaccine efficacy data of LAIV in accordance with clinical trial data resulted in a substantial higher NHB. Moreover, we assumed no difference in duration of protection between LAIV and IV, while some evidence suggests that IVs already wane through the season and LAIV may protect in a second season.^51-53^ However, applying a longer duration of protection of LAIV is not expected to have significant impact on our outcomes, because the analysis assumes LAIV to be given on an annual basis.

No cross-reactivity between influenza subtypes was considered in the analysis, while in reality such mechanisms could exist. For instance, the cross-protection of TIV against the mismatched influenza B lineage has been estimated at 70% of the efficacy against the matched influenza B lineage.^26,54^ If true, our study may overestimate the additional effectiveness of Q-LAIV over TIV.

We used influenza-associated mortality rates that were regressed against respiratory diagnoses, while an ecological study that used all-cause mortality data found substantially higher influenza mortality rates.^55^ However, the use of all-cause mortality data could also result in an overestimation of the number of deaths attributed to influenza, and the use of respiratory diagnoses reflects a conservative approach.

Furthermore, assumptions had to be made for the vaccine prices of Q-LAIV and QIV, as tendered prices for the Dutch setting are unavailable. However, our sensitivity analyses demonstrate that paediatric vaccination is expected to be cost-effective even at substantially higher vaccine prices.

A recent study by Backer et al.^48^ estimated that adding Q-LAIV for 2- to 16-year-olds at 40% coverage would reduce the overall infection attack rate in the Netherlands by 15%, which is lower than the 28% reduction we estimated for adding TIV for 2- to 17-year-olds at 50% coverage. Next to the targeted age range and assumed coverage, this difference may be explained by differences in the model structure and the use of an on average higher vaccine efficacy and longer duration of vaccine protection as compared to those used by Backer et al. Our finding that paediatric vaccination is cost-effective from a healthcare payer’s perspective and cost-saving from a societal perspective is in line with published studies from surrounding European countries.^56-58^

Results of this study are relevant for policy-makers deciding whether to introduce paediatric influenza vaccination in the Netherlands or elsewhere. However, cost-effectiveness is not the only criterion involved in this decision.^59^ For instance, acceptability of the vaccination programme is also important. Although paediatric vaccination was also cost-effective for children themselves, most of its benefits are among other age groups via indirect protection. A non-uniform distribution of advantages of a vaccination programme may well be perfectly acceptable when adverse events of vaccination are mild and public health in general is substantially improved.^18^ However, acceptance among the parents of the children is also important. A recent Dutch survey showed that the willingness to vaccinate their children against influenza was 15%, which was substantially lower than other currently unimplemented vaccines against rotavirus, varicella, and *Meningococcus B*.^60^

Finally, the impact of routine influenza vaccination in early childhood on the long-term development of immunity against influenza viruses is a matter of debate. Accumulating evidence suggests that the first influenza infections in life influence the immune response against subsequent infections (imprinting).^61,62^ However, it is unknown whether vaccination interferes with or promotes imprinting.^61^ LAIV is thought to be a more appropriate vaccine candidate than IV for children naïve to influenza infections because it mimics a natural infection in the upper respiratory tract that activates mucosal antibodies and cross-protective cytotoxic T-cell lymphocytes as well as strain-specific serum antibodies.^63^

## 5. Conclusion

Modelling indicates that paediatric influenza vaccination reduces the influenza morbidity and mortality substantially and is cost-saving from a societal perspective. The highest NHB is observed for vaccination of 2- to 17-year-olds with Q-LAIV and elderly and risk groups with QIV. It is anticipated that results of this study will be useful for policy-makers in deciding whether to introduce paediatric influenza vaccination.

## Data Availability

All relevant data are within the paper. Further details are available from the first author on reasonable request.

## Acknowledgements

This study was sponsored by AstraZeneca. The funding source had no involvement in the study design or collection, analysis, and interpretation of data. The PhD positions of PTdB and FCKD at the University of Groningen have been supported by grants from various pharmaceutical companies, including those developing, producing and marketing influenza vaccines. PTdB is currently employed at the Dutch National Institute for Public Health and the Environment (RIVM); this work is not on behalf of the RIVM. JCW and MJP have received grants and honoraria from various pharmaceutical companies, including those developing, producing and marketing influenza vaccines. RP has participated as a member of AstraZeneca advisory boards and received funding for research projects.

## References

1. World Health Organization. Influenza (Seasonal) 2018 http://www.who.int/news-room/fact-sheets/detail/influenza-(seasonal). Accessed at 1 August 2018.

2. Peasah SK, Azziz-Baumgartner E, Breese J, Meltzer MI, Widdowson MA. Influenza cost and cost-effectiveness studies globally--a review. Vaccine 2013, 31(46):5339–5348.

3. Heikkinen T, Tsolia M, Finn A. Vaccination of healthy children against seasonal influenza: a European perspective. Pediatr Infect Dis J 2013, 32(8):881–888.

4. Poehling KA, Edwards KM, Griffin MR et al. The burden of influenza in young children, 2004-2009. Pediatrics 2013, 131(2):207–216.

5. Antonova EN, Rycroft CE, Ambrose CS, Heikkinen T, Principi N. Burden of paediatric influenza in Western Europe: a systematic review. BMC Public Health 2012, 12:968.

6. Heikkinen T. Influenza in children. Acta Paediatr 2006, 95(7):778–784.

7. Ghendon YZ, Kaira AN, Elshina GA. The effect of mass influenza immunization in children on the morbidity of the unvaccinated elderly. Epidemiol Infect 2006, 134(1):71–78.

8. Charu V, Viboud C, Simonsen L et al. Influenza-related mortality trends in Japanese and American seniors: evidence for the indirect mortality benefits of vaccinating schoolchildren. PLoS One 2011, 6(11):e26282.

9. Vynnycky E, Pitman R, Siddiqui R, Gay N, Edmunds WJ. Estimating the impact of childhood influenza vaccination programmes in England and Wales. Vaccine 2008, 26(41):5321–5330.

10. Pitman RJ, White LJ, Sculpher M. Estimating the clinical impact of introducing paediatric influenza vaccination in England and Wales. Vaccine 2012, 30(6):1208–1224.

11. Baguelin M, Flasche S, Camacho A, Demiris N, Miller E, Edmunds WJ. Assessing optimal target populations for influenza vaccination programmes: an evidence synthesis and modelling study. PLoS Med 2013, 10(10):e1001527.

12. Eichner M, Schwehm M, Eichner L, Gerlier L. Direct and indirect effects of influenza vaccination. BMC Infect Dis 2017, 17(1):308.

13. Ortiz JR, Perut M, Dumolard L et al. A global review of national influenza immunization policies: Analysis of the 2014 WHO/UNICEF Joint Reporting Form on immunization. Vaccine 2016, 34(45):5400–5405.

14. Department of Health. The flu immunisation programme 2013/14 – extension to children 2013 https://assets.publishing.service.gov.uk/government/uploads/system/uploads/attachment_data/file/225360/Children_s_flu_letter_2013.pdf. Accessed at 1 Aug 2018.

15. National Institute for Public Health and the Environment. Flu and flu shot 2015 https://www.rivm.nl/Onderwerpen/G/Griep. Accessed at 1 August 2018.

16. Health Council of the Netherlands. Griepvaccinatie: herziening van de indicatiestelling [In Dutch] 2007 https://www.gezondheidsraad.nl/nl/taak-werkwijze/werkterrein/preventie/griepvaccinatie-herziening-van-de-indicatiestelling. Accessed at 1 August 2018.

17. Health Council of the Netherlands. Werkagenda advisering Vaccinaties Gezondheidsraad 2018 - 2020 [In Dutch] 2018 https://www.rijksoverheid.nl/binaries/rijksoverheid/documenten/brieven/2017/01/31/werkagenda-advisering-vaccinaties-gezondheidsraad/werkagenda-advisering-vaccinaties-gezondheidsraad.pdf. Accessed at 1 August 2018.

18. Houweling H, Verweij M, Ruitenberg EJ, National Immunisation Programme Review Committee of the Health Council of the N. Criteria for inclusion of vaccinations in public programmes. Vaccine 2010, 28(17):2924–2931.

19. Statistics Netherlands. Population; sex, age and marital status, 1 January 2010 https://statline.cbs.nl/Statweb/publication/?DM=SLEN&PA=7461eng&D1=0&D2=0&D3=0-100&D4=60&LA=EN&HDR=T,G3&STB=G1,G2&VW=T. Accessed at 1 Jan 2015.

20. Statistics Netherlands. Life expectancy; gender, age (per year and period of five years) 2010 https://statline.cbs.nl/Statweb/publication/?DM=SLNL&PA=37360ned&D1=0&D2=0&D3=a&D4=89&HDR=G1,T&STB=G2,G3&VW=T. Accessed at 1 Jan 2015.

21. Statistics Netherlands. Birth; key figures 2010 https://statline.cbs.nl/Statweb/publication/?DM=SLEN&PA=37422eng&D1=0-2&D2=60&LA=EN&HDR=T&STB=G1&VW=T. Accessed at 1 Jan 2015.

22. Mossong J, Hens N, Jit M et al. Social contacts and mixing patterns relevant to the spread of infectious diseases. PLoS Med 2008, 5(3):e74.

23. Dolk FCK, De Boer PT, Nagy L et al. Consultations for influenza-like illness influenza-like illness in primary care in The Netherlands; a regression approach. Submitted.

24. World Health Organization. Influenza vaccine viruses and reagents 2015 https://www.who.int/influenza/vaccines/virus/en/. Accessed at 1 Jan 2016.

25. National Institute for Public Health and the Environment. Monitoring vaccination coverage national program for flu prevention [In Dutch] 2015 https://www.rivm.nl/monitoring-vaccinatiegraad-nationaal-programma-grieppreventie-npg. Accessed at 1 Jan 2016.

26. DiazGranados CA, Denis M, Plotkin S. Seasonal influenza vaccine efficacy and its determinants in children and non-elderly adults: a systematic review with meta-analyses of controlled trials. Vaccine 2012, 31(1):49–57.

27. Rivetti D, Jefferson T, Thomas R et al. Vaccines for preventing influenza in the elderly. Cochrane Database Syst Rev 2006(3):CD004876.

28. National Health Care Institute. Guideline for economic evaluations in healthcare 2016 https://english.zorginstituutnederland.nl/publications/reports/2016/06/16/guideline-for-economic-evaluations-in-healthcare. Accessed at Dec 1 2017.

29. Chung JR, Flannery B, Ambrose CS et al. Live Attenuated and Inactivated Influenza Vaccine Effectiveness. Pediatrics 2019, 143(2).

30. Buchan SA, Booth S, Scott AN et al. Effectiveness of Live Attenuated vs Inactivated Influenza Vaccines in Children During the 2012-2013 Through 2015-2016 Influenza Seasons in Alberta, Canada: A Canadian Immunization Research Network (CIRN) Study. JAMA Pediatr 2018, 172(9):e181514.

31. Caspard H, Mallory RM, Yu J, Ambrose CS. Live-Attenuated Influenza Vaccine Effectiveness in Children From 2009 to 2015-2016: A Systematic Review and Meta-Analysis. Open Forum Infect Dis 2017, 4(3):ofx111.

32. GOV.UK. Seasonal flu vaccine uptake in children of primary school age: monthly data, 2017 to 2018 2018 https://www.gov.uk/government/statistics/seasonal-flu-vaccine-uptake-in-children-of-primary-school-age-monthly-data-2017-to-2018. Accessed at 1 August 2018.

33. Carrat F, Vergu E, Ferguson NM et al. Time lines of infection and disease in human influenza: a review of volunteer challenge studies. Am J Epidemiol 2008, 167(7):775–785.

34. van den Wijngaard CC, van Asten L, Meijer A et al. Detection of excess influenza severity: associating respiratory hospitalization and mortality data with reports of influenza-like illness by primary care physicians. Am J Public Health 2010, 100(11):2248–2254.

35. van den Wijngaard CC, van Asten L, Koopmans MP et al. Comparing pandemic to seasonal influenza mortality: moderate impact overall but high mortality in young children. PLoS One 2012, 7(2):e31197.

36. Statistics Netherlands. Consumer prices; price index 2015=100 2017 http://statline.cbs.nl/Statweb/publication/?DM=SLEN&PA=83131ENG&D1=0&D2=0&D3=64,77,90,103,116,129,142,155,168,181,194,207,220,233,246,259,272,285&LA=EN&HDR=T&STB=G1,G2&VW=T. Accessed at Dec 1 2017.

37. Stichting Nationaal Programma Grieppreventie (SNPG). News letter SNPG November 2017 for health-care organizations (In Dutch) 2017 https://www.snpg.nl/2017/11/23/nieuwsbrief-snpg-november-2017-zorgorganisaties/. Accessed at 1 Aug 2018.

38. Mangen MJ, Rozenbaum MH, Huijts SM et al. Cost-effectiveness of adult pneumococcal conjugate vaccination in the Netherlands. Eur Respir J 2015, 46(5):1407–1416.

39. Versteegh MM, Vermeulen KM, Evers SMAA, de Wit GA, Prenger R, Stolk EA. Dutch Tariff for the Five-Level Version of EQ-5D. Value Health 2016, 19(4):343–352.

40. Statistics Netherlands. Projected Demograpich Development, 2015-2060 [In Dutch] 2014 https://statline.cbs.nl/Statweb/publication/?DM=SLNL&PA=83224NED&D1=a&D2=0,5,10,15,20,25,30,35,40,l&VW=T. Accessed at 1 Jul 2016.

41. National Health Care Institute. Cost-effectiveness in practice [In Dutch] 2015 https://www.zorginstituutnederland.nl/publicaties/rapport/2015/06/26/kosteneffectiviteit-in-de-praktijk. Accessed at 1 Dec 2018.

42. Barton GR, Briggs AH, Fenwick EA. Optimal cost-effectiveness decisions: the role of the cost-effectiveness acceptability curve (CEAC), the cost-effectiveness acceptability frontier (CEAF), and the expected value of perfection information (EVPI). Value Health 2008, 11(5):886–897.

43. Osterholm MT, Kelley NS, Sommer A, Belongia EA. Efficacy and effectiveness of influenza vaccines: a systematic review and meta-analysis. Lancet Infect Dis 2012, 12(1):36–44.

44. Ministry of Health Welfare and Sport. Letter to the parlement on flu measures [In Dutch] 2018 https://www.rijksoverheid.nl/binaries/rijksoverheid/documenten/kamerstukken/2018/10/10/kamerbrief-over-maatregelen-griep/kamerbrief-over-maatregelen-griep.pdf. Accessed at 15 Jan 2019.

45. Pitman RJ, Nagy LD, Sculpher MJ. Cost-effectiveness of childhood influenza vaccination in England and Wales: Results from a dynamic transmission model. Vaccine 2013, 31(6):927–942.

46. Pitman R, Fisman D, Zaric GS et al. Dynamic transmission modeling: a report of the ISPOR-SMDM Modeling Good Research Practices Task Force--5. Value Health 2012, 15(6):828–834.

47. Newall AT, Chaiyakunapruk N, Lambach P, Hutubessy RCW. WHO guide on the economic evaluation of influenza vaccination. Influenza Other Respir Viruses 2018, 12(2):211–219.

48. Backer JA, van Boven M, van der Hoek W, Wallinga J. Vaccinating children against influenza increases variability in epidemic size. Epidemics 2019, 26:95–103.

49. Woolthuis RG, Wallinga J, van Boven M. Variation in loss of immunity shapes influenza epidemics and the impact of vaccination. BMC Infect Dis 2017, 17(1):632.

50. National Institute for Public Health and the Environment. Influenza vaccination in the Netherlands. Background information for the Health Council of the Netherlands 2019 https://www.rivm.nl/bibliotheek/rapporten/2019-0002.pdf. Accessed at 25 July 2019.

51. Ferdinands JM, Fry AM, Reynolds S et al. Intraseason waning of influenza vaccine protection: Evidence from the US Influenza Vaccine Effectiveness Network, 2011-12 through 2014-15. Clin Infect Dis 2017, 64(5):544–550.

52. Kissling E, Nunes B, Robertson C et al. I-MOVE multicentre case-control study 2010/11 to 2014/15: Is there within-season waning of influenza type/subtype vaccine effectiveness with increasing time since vaccination? Euro Surveill 2016, 21(16).

53. Tam JS, Capeding MR, Lum LC et al. Efficacy and safety of a live attenuated, cold-adapted influenza vaccine, trivalent against culture-confirmed influenza in young children in Asia. Pediatr Infect Dis J 2007, 26(7):619–628.

54. Tricco AC, Chit A, Soobiah C et al. Comparing influenza vaccine efficacy against mismatched and matched strains: a systematic review and meta-analysis. BMC Med 2013, 11:153.

55. McDonald SA, van Wijhe M, van Asten L, van der Hoek W, Wallinga J. Years of life lost due to influenza-attributable mortality in older adults in the Netherlands: a competing risks approach. Am J Epidemiol 2018, 187(8):1791–1798.

56. Damm O, Eichner M, Rose MA et al. Public health impact and cost-effectiveness of intranasal live attenuated influenza vaccination of children in Germany. Eur J Health Econ 2015, 16(5):471–488.

57. Baguelin M, Camacho A, Flasche S, Edmunds WJ. Extending the elderly- and risk-group programme of vaccination against seasonal influenza in England and Wales: a cost-effectiveness study. BMC Med 2015, 13:236.

58. Thorrington D, Jit M, Eames K. Targeted vaccination in healthy school children - Can primary school vaccination alone control influenza? Vaccine 2015, 33(41):5415–5424.

59. Ahout I, Ferwerda G, de Groot R. Influenza vaccination in kids, are you kidding me? J Infect 2014, 68 Suppl 1:S100–107.

60. van Lier A, Ferreira JA, Mollema L, Sanders EAM, de Melker HE. Intention to vaccinate universally against varicella, rotavirus gastroenteritis, meningococcal B disease and seasonal influenza among parents in the Netherlands: an internet survey. BMC Res Notes 2017, 10(1):672.

61. Viboud C, Epstein SL. First flu is forever. Science 2016, 354(6313):706–707.

62. Monto AS, Malosh RE, Petrie JG, Martin ET. The Doctrine of Original Antigenic Sin: Separating Good From Evil. J Infect Dis 2017, 215(12):1782–1788.

63. Mohn KG, Smith I, Sjursen H, Cox RJ. Immune responses after live attenuated influenza vaccination. Hum Vaccin Immunother 2018, 14(3):571–578.

